# Non-communicable diseases Prevalence by HIV Status in East Africa: A protocol for a systematic review and Meta-analysis

**DOI:** 10.1101/2023.12.05.23299514

**Authors:** Asani Kasango, Alex Daama, Lilian Negesa

## Abstract

**Introduction:** Over the past decade, HIV care and treatment programs have grown exponentially, resulting in improved survival among persons living with HIV. In the same period, the prevalence of non-communicable diseases has been reported to be higher among people living with HIV as compared to their HIV-negative counterparts. In this review, we aim to assess the prevalence of non-communicable diseases stratified by HIV status in East Africa.

**Methods:** We plan to assess observational studies especially case-control, cohort, and cross-sectional studies and clinical trials. These studies will be related to non-communicable diseases and done in East Africa. This review will be for studies done in East Africa from 1^st^ Jan 2013 to 31^st^ Dec 2023. The team will search databases such as East African Medical Journal, PubMed, Pan African Medical Journal, Scopus, African Health Sciences Journal, African Index, Medicus, and African Journals Online among others. We will use subheadings such as HIV, NCDs, and East Africa to search for relevant Journals. The I^2^ will be used to assess the heterogeneity of individual studies. All statistical analyses will be performed in STATA V15, and results will be graphically presented on a forest plot.

**Discussion:** With the rising burden of non-communicable diseases globally, there is much to learn whether the prevalence differs by HIV status or not.

## Introduction

HIV/AIDS is a priority public health concern that has existed for over 30 years. It has contributed to 75 million cases and 32 million deaths (1). There has been a rapid increase in coverage of HIV care and treatment within the past decade (2). These programs have successfully controlled viremia and Acquired Immunodeficiency syndrome through ARVs (3). People living with HIV (PLHIV) are now aging and are now susceptible to non-communicable diseases (NCDs) (3,4).

NCDs account for 70% of global deaths per annum and since a significant proportion of PLHIV are aging, it’s predicted that NCDs will become the leading cause of morbidity and mortality among PLHIV by 2030 (5). Non-communicable diseases are associated with increased morbidity and mortality in the general population and among PLHIV. For instance, a report from the global burden of diseases, injuries, and Risk Factor (GBD) study indicated a 40% increase in NCDs disability-adjusted life years (DALYs) between 2007 and 2017, and NCDs alone contributed to 62% of DALYs in the same period (6). The growing burden of NCDs has necessitated the need to compare the NCD prevalence by HIV status to inform program managers and Health policymakers in East Africa to adopt, implement, and scale up appropriate interventions for most at-risk subpopulations for NCD screening and management.

### Aim

This review aims to systematically compare the prevalence of NCDs by HIV status in East Africa.

## Methods

### Study design

The Preferred Reporting Items for Systematic Reviews and Meta-analyses (PRISMA) guidelines (7) will be followed during the conduct of this systematic review and Meta-Analysis.

### Study setting

The setting for the study will be studies on NCDs done from 1^st^ Jan 2013-31^st^ Dec 2023 in East Africa.

### Population

Studies that assessed any NCDs will be included.

### Exposures of Interest

Exposures of interest will be HIV, ART use, and traditional risk factors for NCDs such as overweight, obesity, alcohol use, smoking, excessive salt consumption, and unhealthy diets.

### Comparison

We will compare NCD prevalence by HIV status.

### Outcome

The outcome will be the prevalence of NCDs. The non-communicable diseases to be considered include hypertension (HT), type 2 diabetes (T2DM), mental illnesses, other coronary heart diseases, cancer, and kidney disease will be assessed.

### Time frame

Studies on NCDs published from 1^st^ January 2013 to 31^st^ Dec 2023.

### Search strategy

A comprehensive search for eligible studies will be done using databases such as East African Medical Journal, PubMed, Pan African Medical Journal, Scopus, African Health Sciences Journal, African Index, Medicus, African Journals Online, and WHO databases among others. We will use subject headings to elicit studies. Furthermore, key terms will include HIV, NCDs, prevalence, and East Africa.

### Inclusion criteria

The studies will be selected for inclusion using the PECOTS framework where; P is Population, E is exposure, C is Comparison, O is Outcome, T is Time frame and S is Study setting (8). We will additionally include cross-sectional, case-control, cohort studies, and clinical trials. Furthermore, only studies reported in English will be included in this review.

### Exclusion criteria

We will exclude non-empirical, qualitative, and epidemiological studies that do not report measures of association including prevalence, incidence, odds ratios (ORs), and Hazard ratios (HRs).

### Study selection

AK and LN will search for eligible studies. We will screen the title and abstract of retrieved articles using Microsoft Excel. The selected articles will be retrieved for data abstraction. Any disagreements that will emerge during data abstraction will be addressed by consulting the third author (AD). In case of unclear and or missing data, the corresponding authors of the primary studies will be emailed. If he/she does not respond after two weeks, he/she will be telephone contacted (if a phone is available and or reachable) and an email reminder will be sent. If no response is obtained after 1 week, the article will be removed and will be reported in the PRISMA flow diagram.

### Quality assessment

To ensure quality assessment, the Newcastle-Ottawa Scale (NOS) will be used. A ‘star’ structure will be used to score and assess the quality of studies. The maximum score for any study is 8. Studies with a score ≥5 or higher will be considered high quality.

### Data extraction

The following data will be extracted: author’s details, participants’ characteristics including their sociodemographic attributes, the year of article publication, study type, study setting, HIV status, study sample size, ART status, prevalence of NCDs, overweight, obesity and other risk factors for NCDs.

### Statistical Analysis

Data analysis will be performed using STATA V15.0. Furthermore, random-effects meta-analysis will be employed to account for variability in the prevalence of different studies. We will use a binomial distribution model to predict prevalence estimates. We will then apply the Freeman-Turkey double arcsine to transform parameters. After normalizing the estimates, pooled estimates will be calculated.

The heterogeneity of studies will be assessed using Cochran’s Q statistic (and this will be expressed using X^2^ and p-value). Additionally, we will perform the I^2^ statistic. The sources of heterogeneity will be explored through subgroup analyses and by performing meta-regression analysis. The study design, sample size, methodology quality, and the year of publication will be assessed using univariate meta-regression models. Sensitivity analysis will also be performed to investigate if the results are influenced by a single study. We will present pooled estimates to account for heterogeneity.

### Anticipating limitations and bias

The authors recognize that there will be limitations to this review. The major limitation of this review is publication bias. This will be ascertained by conducting a sensitivity analysis. By doing this, we will determine each study status by how it graphically fits on the funnel plot. On the funnel plot, studies with high precision tend to plot closer to the meanwhile those whose precision is low disperse away from the mean. We will further use Egger and Begg’s test to ascertain publication bias. Any p-value < 0.1 will indicate publication bias.

## Discussion

We plan to summarize the totality of evidence on the prevalence of NCDs by HIV status in East Africa. Findings may in part be useful in identifying further research priorities to address these gaps and to support a better understanding of the burden of NCDs in the region.

### Strengths and Limitations

This will be a meta-analysis of studies done in East Africa and will provide a better prevalence of NCDs by HIV status. It is also one of the few reviews in East Africa to stratify NCD prevalence by HIV status, making its results more generalizable compared to individual studies. The key limitation of this review is that it will only include studies done in English. Furthermore, this review will be limited by publication bias.

## Conclusion

In conclusion, a review such as this could help to inform the world about the prevalence of non-communicable diseases by HIV status in Africa.

## Data Availability

N/A

## Declarations

### Ethics approval and consent to participate

Not applicable

### Consent for publication

Not Applicable

### Availability of Data and Methods

Data sharing does not apply to this article as no datasets were generated or analyzed during the current study.

### Compelling interests

We declare no conflict of interest.

### Authors’ contributions

AK conducted the pre-research, designed the review, developed the search strategy, and drafted the manuscript; LN provided input on the design, developed the search strategy, and edited the manuscript; All authors reviewed and approved the final manuscript.

## Acknowledgements

We acknowledge the principal investigators for the primary studies for sacrificing resources, time, and effort for the different studies. We equally acknowledge the research assistants and study participants in the primary studies.

